# Antibodies to SARS-CoV-2 and risk of future sickness

**DOI:** 10.1101/2020.09.14.20194308

**Authors:** Joakim Dillner, K. Miriam Elfström, Jonas Blomqvist, Carina Eklund, Camilla Lagheden, Sara Nordqvist-Kleppe, Cecilia Hellström, Jennie Olofsson, Eni Andersson, August Jernbom Falk, Sofia Bergström, Emilie Hultin, Elisa Pin, Anna Månberg, Peter Nilsson, My Hedhammar, Sophia Hober, Johan Mattsson, Laila Sara Arroyo Mühr, Kalle Conneryd Lundgren

**Affiliations:** Karolinska University Laboratory, Karolinska University Hospital, SE-141 86 Stockholm, Sweden.; Division of Affinity Proteomics, Department of Protein Science, KTH Royal Institute of Technology, SciLifeLab, SE-171 65 Stockholm, Sweden; Division of Protein Technology, Department of Protein Science, KTH Royal Institute of Technology, Albanova, SE-144 21 Stockholm, Sweden; Karolinska University Hospital, SE-141 86 Stockholm, Sweden

**Keywords:** SARS-CoV-2, Corona, Coronavirus, antibodies, sick leave, healthcare workers

## Abstract

**Background:** The extent that antibodies to SARS-CoV-2 may protect against future virus-associated disease is unknown.

**Method:** We analyzed 12928 healthy hospital employees for SARS-CoV-2 antibodies and compared results to participant sick leave records (Clinical trial registration: https:ClinicalTrials.gov NCT04411576).

**Results:** Subjects with viral serum antibodies were not at excess risk for future sick leave (Odds Ratio (OR): 0.85 (95% Confidence Interval (CI) (0.85 (0.43-1.68)). By contrast, subjects with antibodies had an excess risk for sick leave in the past weeks (OR: 3.34 (2.98-3.74)).

**Conclusion:** Presence of viral antibodies marks past disease and protection against excess risk of future disease.

## Background

To design strategies for SARS-CoV-2 control, knowledge of whether exposed individuals are immune against future disease is critical [1]. The incubation time from exposure to onset of symptoms has been estimated to last a median of six days, with peak infectiousness occurring zero to two days before onset of symptoms [2] and pre-symptomatic spread is estimated to account for a substantial proportion of disease transmission [2,3]. Infectiousness decreases with increasing time after onset of symptoms [4] and some individuals may remain asymptomatic despite being virus positive [5]. The IgG response develops rather slowly, commonly concomitantly with symptom resolution and increases in subsequent weeks. One report found that all COVID-19 patients had become seropositive 19 days after onset of symptoms [6]. Although there are many studies on viral antibodies and immunity, it is still uncertain to what extent immunity exists and what the predictive value of presence of viral antibodies is. A problem is that studies that are based on past sickness are fraught with recall biases (subjects knowing their antibody status preferentially recalling events) and that prospective studies using future sickness as endpoint need to be very large.

## Methods

The Karolinska University Hospital is one of the largest university hospitals in Europe, with about 15,300 employees (employment during enrollment, 23rd April - 22nd May 2020). The hospital announced that all employees who were at work (without ongoing sickness) were welcome to participate in a study on SARS-CoV-2. We enrolled 14,057 participants who all signed a written informed consent that also included permission to extract data from the employer’s administrative databases that contain data on sick leave. Most participants were between 30-59 years old and 79% were females (Supplementary Table 1). Analysis results were reported to participants, but analyses took 4 weeks post sampling to complete. Sick leave data was obtained from 6 weeks before sampling until 2 weeks after sampling. The study was approved by the National Ethical Review Agency of Sweden (Decision number 2020–01620). Trial registration number: https:ClinicalTrials.gov NCT04411576

### Analyses of SARS-CoV-2 antibodies

Whole blood was collected in serum-separating tubes and centrifuged under 2000 x g for ten minutes. Serum samples were inactivated by heating to 56 degrees Celsius for 30 minutes and then stored at minus 20 degrees Celsius until further analysis.

Different SARS-CoV-2 protein constructs and different production hosts were compared for expression of viral proteins using the mammalian HEK cell line as starting point. The evaluation of different production hosts was based on degree of concordance in antibody reactivity of the alternative hosts with the virus proteins produced in the HEK cells. Thereafter, the most efficient production and purification pipeline was chosen. Consequently, antigen reactivity was measured towards three different virus protein-variants, (i) Spike trimers comprising the prefusion-stabilized spike glycoprotein ectodomain [7] expressed in HEK cells and purified using a C-terminal Strep II tag), (ii) Spike S1 domain, expressed in CHO cells and purified using C- terminal HPC4-tag, and (iii) Nucleocapsid protein, expressed in E.coli and purified using a C-terminal His-tag.

The sera were analyzed using a multiplex antigen bead array in high throughput 384-plates format using a FlexMap3D instrument (Luminex Corp) with IgG detection [8]. The cut-off for seropositivity was for each antigen defined as mean +6SD of 12 negative control samples included in each analysis batch. To be assigned as IgG positive, a sample was required to show reactivity against at least two of the three included viral antigens. Serum IgG bound to antigen coated beads was detected by F(ab^’^)2-Goat anti-Human IgG Fc Secondary Antibody, PEfluorescent anti-hIgG (Invitrogen, H10104. Validation procedure is described at https://www.thermofisher.com/se/en/home/life-science/antibodies/invitrogen-antibody-validation.html) and recorded as relative fluorescence intensity (AU). Four positive controls were re-run on every assay-plate and had a mean inter-assay coefficient of variation of 10.1% (8.0-13.3%), based on absolute intensity levels.

The serology assay was evaluated based on the analyses of 243 samples from Covid-19 subjects (defined as PCR-positive individuals sampled more than 16 days after positive PCR test) and 442 negative control samples (defined as samples collected 2019 or earlier, including 26 individuals with confirmed infections of other Coronaviruses than SARS-CoV-2). Based on these samples, the assay had a 99.2% sensitivity and 99.8% specificity.

### Data analyses

With conventional statistical power and two-sided tests of significance, and assuming a cumulative proportion of sick leave among non-exposed persons of 30% and that 10% of the cohort might be exposed, at least 3800 subjects would need to be enrolled to be able to detect associations of 1.4 or greater, a level which was considered to be medically meaningful. Descriptive statistics examined test results by age, sex and sick leave. A multinomial logistic regression model was used to examine the association between test results and sick leave measured as a categorical variable. The final model was adjusted for age in ten-year categories and gender. For subjects with sick leave in more than one category, the period with the most days on sick leave was chosen. If two periods had an equal number of sick leave days, the period further back in time was chosen. Analyses were completed using SAS 9.4, Cary, NC.

## Results

We invited all employees currently at work at a large hospital in Sweden to participate in a longitudinal cohort study of SARS-CoV-2 testing in relation to both past and future sick leave. We enrolled 14,052 participants (Figure 1). After exclusion of participants not formally employed (e.g. medical students) and subjects with invalid tests, the final cohort consisted of 12,928 subjects with complete data on sick leave and valid SARS-CoV-2 antibody results (Figure 1).

**Figure 1.**
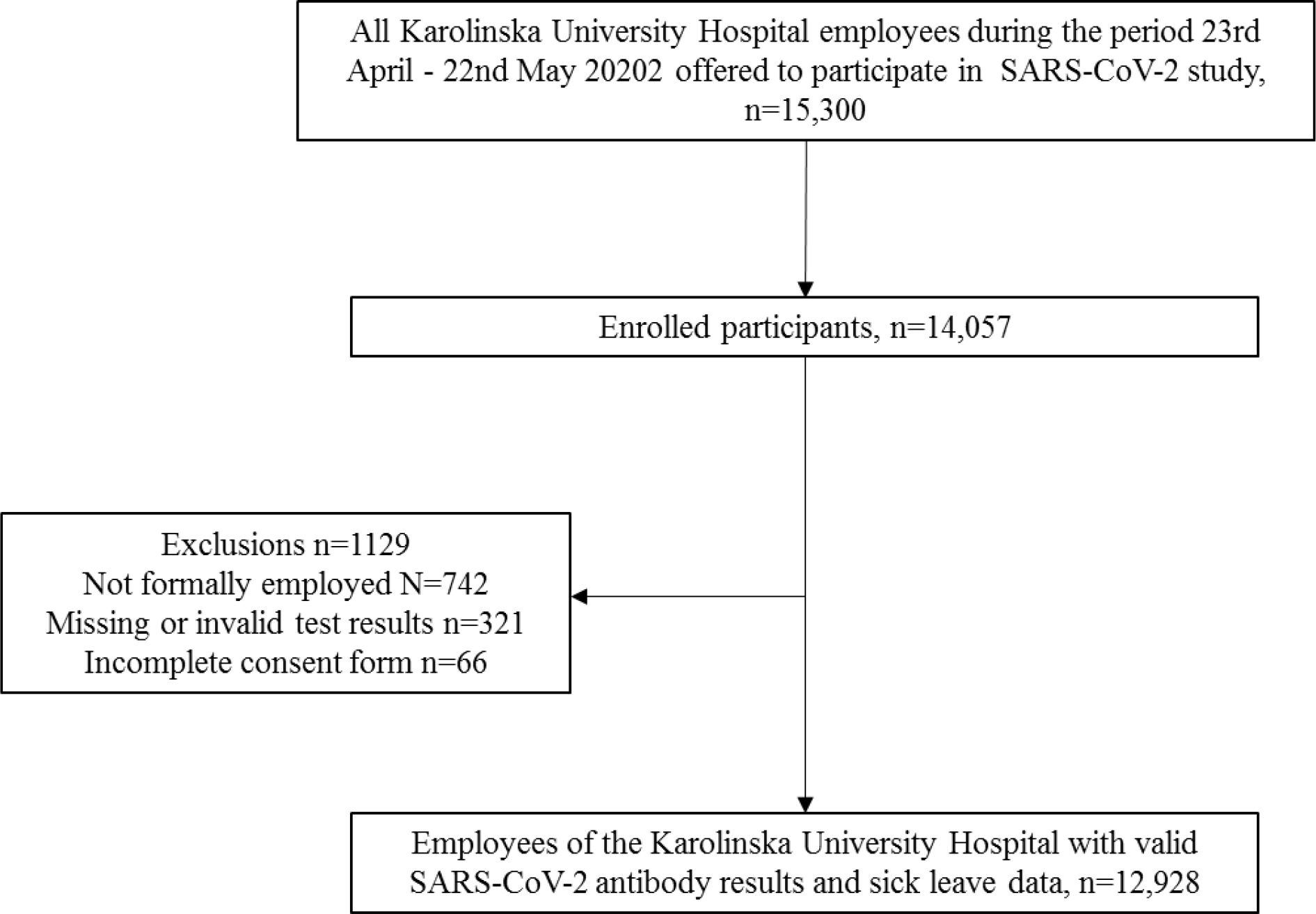
STROBE flowchart of study participants

The overall number and proportion of employees that tested positive in the antibody tests are shown by age in ten-year spans in Table 1. The proportion of serology-positive subjects was greatest among the youngest employees and decreased significantly with increasing age (p-value for trend < 0.0001).

**Table 1.** Detection of antibodies to the SARS CoV-2 virus among 12928 employees of the Karolinska University Hospital, by age

| Age | Serology positive |  | Total |
| --- | --- | --- | --- |
|  | n | % (95% CI)<sup>a</sup> |  |
| <29 | 249 | 16.4 (14.6-18.3) | 1 522 |
| 30-39 | 383 | 12.1 (11.0-13.3) | 3 172 |
| 40-49 | 370 | 11.4 (10.4-12.6) | 3 238 |
| 50-59 | 313 | 10.2 (9.2-11.3) | 3 066 |
| 60+ | 166 | 8.6 (7.4-9.9) | 1 930 |
| Total | 1 481 | 11.5 (10.9-11.8) | 12 928 |

<sup>a</sup>Cochran-Armitage Trend Test for decreasing prevalence with increasing age, p-value <0.0001

Positivity in serology was significantly associated with an excess risk for having been on sick leave in the past 6 weeks (Table 2) but did not confer any excess risk for future sick leave for the coming two weeks after testing (Table 2). The mutual adjustments (age, sex and serostatus) in the multivariate model had only minor effects on the estimates (Supplementary Table 2).

**Table 2.**
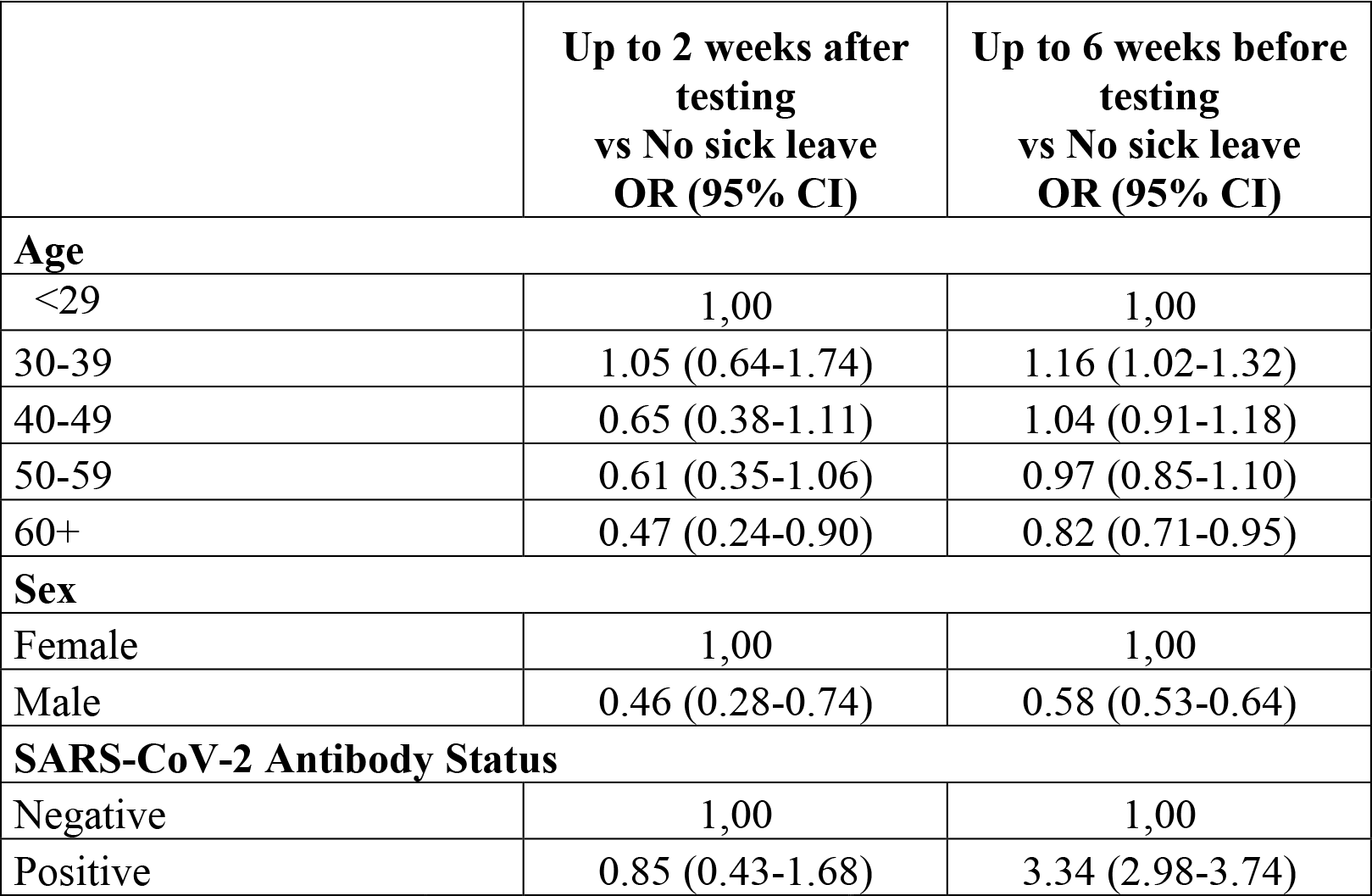
Association between covariates and sick leave, mutually adjusted

|  | **Up to 2 weeks after**<br>**testing**<br>**vs No sick leave**<br>**OR (95% CI)** | **Up to 6 weeks before**<br>**testing**<br>**vs No sick leave**<br>**OR (95% CI)** |
| --- | --- | --- |
| **Age** |  |  |
| <29 | 1,00 | 1,00 |
| 30-39 | 1.05 (0.64-1.74) | 1.16 (1.02-1.32) |
| 40-49 | 0.65 (0.38-1.11) | 1.04 (0.91-1.18) |
| 50-59 | 0.61 (0.35-1.06) | 0.97 (0.85-1.10) |
| 60+ | 0.47 (0.24-0.90) | 0.82 (0.71-0.95) |
| **Sex** |  |  |
| Female | 1,00 | 1,00 |
| Male | 0.46 (0.28-0.74) | 0.58 (0.53-0.64) |
| **SARS-CoV-2 Antibody Status** |  |  |
| Negative | 1,00 | 1,00 |
| Positive | 0.85 (0.43-1.68) | 3.34 (2.98-3.74) |

As Covid-19 is known to preferentially affect older subjects and have a long sickness duration, we next analyzed the length of sick leave in relation to age and serology status (Table 3). No past sick leave was found for 66% of the antibody-negative subjects, whereas only 35% of the antibody-positive subjects had no past sick leave (Table 3). The association of seropositivity with past sick leave was highly significant (OR: 3.34 (2.98-3.74)) and mostly due to sick leave longer than five days (Table 3). Figure 2 displays the proportion of participants on sick leave in relation to their antibody status. The typical seasonal pattern with reported sick leave declining over time during spring was seen for both seronegative and seropositive subjects (Figure 2), but the elevated excess odds ratio for past sick leave was similar for the different weeks before testing.

**Table 3.** Duration of sick leave in days, by SARS-CoV-2 serology test result and age

| Antibody status | Age | Cumulative number of sick leave days over the period<sup>a</sup> |  |  |  |  |
| --- | --- | --- | --- | --- | --- | --- |
|  |  | 0 days<br>n (%) | 1-5 days<br>n (%) | 6-10 days<br>n (%) | 11-15 days<br>n (%) | ≥16 days<br>n (%) |
| Negative | <29 | 857 (67.3) | 312 (24.5) | 78 (6.1) | 13 (1.0) | 13 (1.0) |
|  | 30-39 | 1,815 (65.1) | 677 (24.3) | 209 (7.5) | 57 (2.0) | 31 (1.1) |
|  | 40-49 | 1,934 (67.4) | 617 (21.5) | 212 (7.4) | 54 (1.9) | 51 (1.8) |
|  | 50-59 | 1,897 (68.9) | 519 (18.9) | 211 (7.7) | 77 (2.8) | 49 (1.8) |
|  | 60+ | 1,282 (72.7) | 287 (16.3) | 119 (6.8) | 46 (2.6) | 30 (1.7) |
|  | **Total n, % (95% CI)** | **7,875, 68.8 (67.9-69.6)** | **2,412, 21.1 (20.3-21.8)** | **829, 7.2 (6.8-7.7)** | **247, 2.2 (1.9-2.4)** | **174, 1.5 (1.3-1.7)** |
| Positive | <29 | 109 (43.8) | 63 (25.3) | 55 (22.1) | 16 (6.4) | 6 (2.4) |
|  | 30-39 | 143 (37.3) | 117 (30.6) | 79 (21.4) | 36 (9.4) | 8 (2.1) |
|  | 40-49 | 143 (38.7) | 97 (26.2) | 79 (21.4) | 42 (11.4) | 9 (2.4) |
|  | 50-59 | 113 (36.1) | 60 (19.2) | 89 (28.4) | 29 (9.3) | 22 (7.0) |
|  | 60+ | 68 (41.0) | 33 (19.9) | 37 (22.3) | 21 (12.7) | 7 (4.2) |
|  | **Total n, % (95% CI)** | **576, 38.9 (36.4-41.4)** | **370, 25.0 (22.9-27.3)** | **339, 22.9 (20.8-25.1)** | **144, 9.7 (8.3-11.3)** | **52, 3.5 (2.7-4.6)** |

<sup>a</sup>Sick leave days in the 6 weeks prior to testing

**Figure 2.**
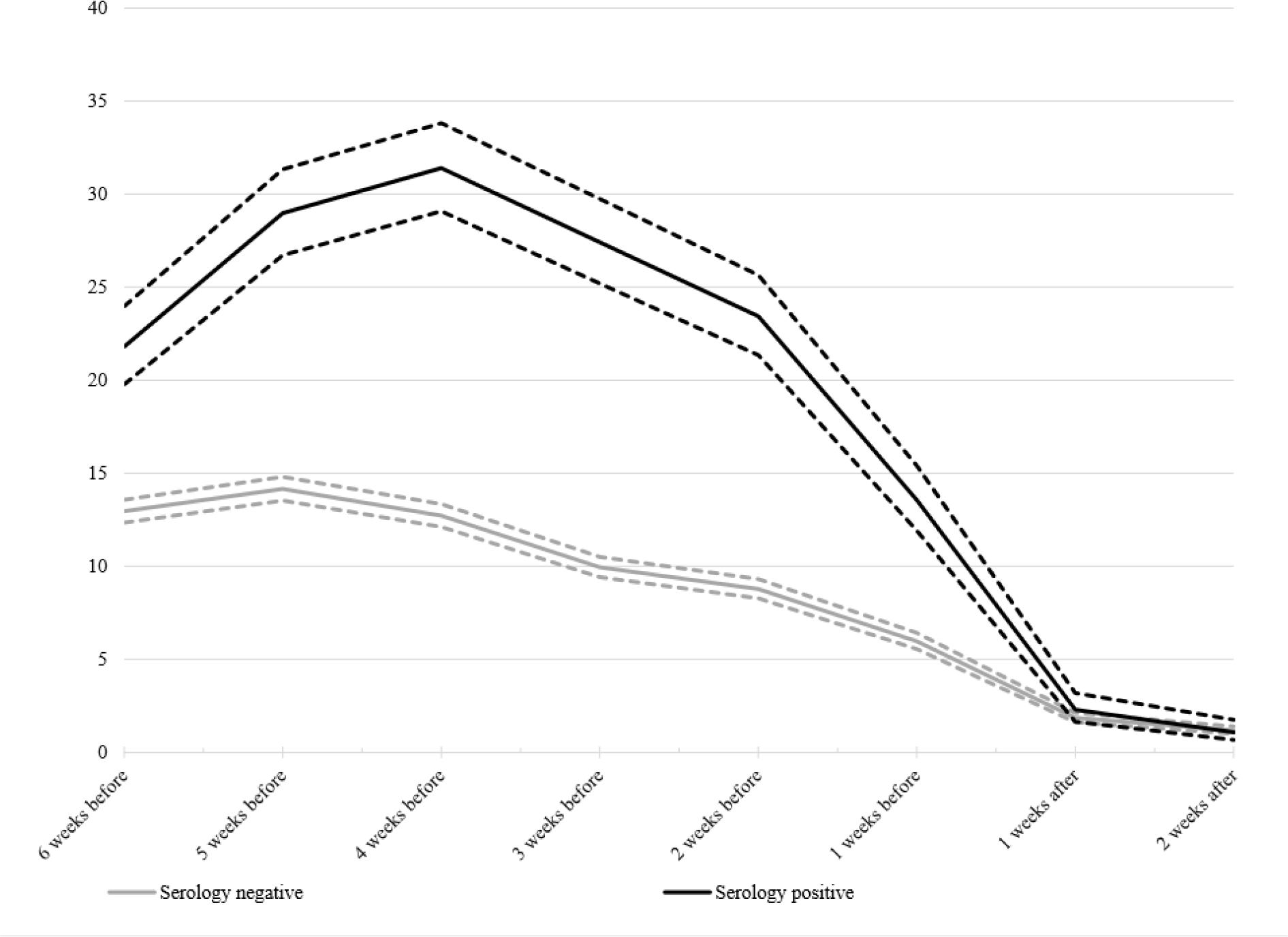
Proportion (%) of healthcare workers on sick leave during the study period, by serology test result-. Dashed lines around the lines denote the 95% confidence intervals.

Similarly, the sick leave curves for seropositive and seronegative subjects are superimposed on each other after testing (Figure 2), in line with the absence of excess risk after testing (Table 2).

## Discussion

We provide large-scale data that antibodies to the SARS-CoV-2 virus associated with protection against excess risk of future sick leave in a large, well-defined cohort of hospital employees. Antibodies were instead associated with past sick leave and serology was thus found to be useful for distinguishing whether subjects who are exposed to the virus may be at risk for development of virus-associated disease or not. Although we find an association of antibodies with protection against excess risk of future disease, it is possible that the antibodies are not directly inducing protection but that their detection SARS-CoV-2 is associated with lowered risk for disease by some indirect association, e.g. if they associate with a resolved disease that has induced cellular immunity.

Strengths of our study include the fact that it was a large and systematically enrolled cohort that used administrative sick leave data and was therefore not hampered by the recall bias to which studies collecting information from participants can be subjected. We used a carefully validated serology platform that contained several SARS-CoV-2 proteins. Test results were delivered to participants, but as the analyses took more than two weeks to complete, registered sick leave after enrollment occurred before the results were delivered and the strong effect seen where serology protected from future excess risk of sickness could thus not have been induced by knowledge of the test results.

A limitation is that we were not able to study infections occurring more than six to seven weeks before enrollment, as community transmission of SARS-CoV-2 in the region started only about six to seven weeks before the study. Participants were not questioned about present or prior symptoms, but the hospital rules were clear that employees with symptoms should not be at work and we had, by design, decided to use only sick leave data to avoid possible recall bias. Hospital rules state that also employees working from home that develop COVID-19 symptoms must report this as sick leave.

We conclude that antibody testing is a powerful tool for identification of subjects who have had prior virus exposure and are protected against future disease. Although it is commonly assumed that antibodies mark immunity, it is important that it has now been shown in a large cohort study that seropositive subjects have no excess risk for future sick leave.

We would like to caution that there is a large number of serology tests currently on the market and the extent of their validation may vary. Also, none of these tests have been specifically validated for the indication proposed here (to separate exposed subjects who are protected from future disease from exposed subjects at risk to develop disease in the future).

In summary, the present study has found that validated antibody testing may be helpful in SARS-CoV-2 screening strategies as antibody-positive subjects were found to have no excess risk for future disease in the weeks after testing.

## Data Availability

The data constitutes sensitive data about health of human research subjects. However, pseudonymised, individual-level data that allow full replication of the results in this article are freely available from or from the Karolinska University Hospital data analysis department

https://www.tableau.karolinska@sll.se

## Acknowledgements

We would like to thank Suyesh Amatya, Helena Andersson, Shaghayegh Bayati, Emine Eken, Pedram Farsi, Yasmin Hussein, Roxana Merino Martinez, Sara Mravinacova, Björn Pfeifer, Ulla Rudsander, Sadaf Sakina Hassan, Ronald Sjöberg, Balazs Szakos, Hanna Tegel, and Emel Yilmaz for excellent technical assistance.

## Author contributions

JD led the study design, data collection, and conceptualization of the analysis in collaboration with KCL. CE, CL, SNK, CH, JO, EA, AJF, SB, EH, EP, AM, PN, MH, SH, LSAM were responsible for preparing and processing samples for laboratory analysis as well as developing and conducting the analyses. JD, KME, JB, and JM analyzed the data. JD and KME wrote the manuscript with support from the co-authors. All authors were involved in the critical revision of the article.

## Notes

**Competing Interests:** None of the authors have any conflicts of interest to declare.

**Funding:**This work was supported by the Karolinska University Hospital (KUH); the County Council of Stockholm; Erling-Persson family foundation; KTH Royal Institute of Technology; Creades and SciLifeLab. Role of funder was as employer of the study team, providing facilities and resources (KUH, KTH and SciLifeLab) and also assisted with enrolment (KUH). There were no other roles for the funders in design or execution of the study, in analysis and interpretation of data or in the decision to submit for publication.

### Competing Interest Statement

The authors have declared no competing interest.

### Clinical Trial

NCT04411576

### Funding Statement

This work was supported by the Karolinska University Hospital (KUH); the County Council of Stockholm; Erling-Persson family foundation; KTH Royal Institute of Technology; Creades and SciLifeLab. Role of funder was as employer of the study team, providing facilities and resources (KUH, KTH and SciLifeLab) and also assisted with enrolment (KUH). There were no other roles for the funders in design or execution of the study, in analysis and interpretation of data or in the decision to submit for publication.

### Author Declarations

The study was approved by the National Ethical Review Agency of Sweden (Decision number 2020-01620).

